# International Consensus Guideline on Management of Genitourinary Adverse Events Associated with Prostate Cancer Radiotherapy

**DOI:** 10.64898/2026.06.14.26355615

**Authors:** Anna M Dornisch, Filippo Alongi, Leslie Ballas, Alison Birtle, Pierre Blanchard, Richard J. Bryant, Ananya Choudhury, Matthew R. Cooperberg, Alan Dal Pra, Jeremy de Leon, Robert T. Dess, Cédric Draulans, William A. Hall, Babbin Sam John, Sophia C. Kamran, Amar U. Kishan, Alastair D. Lamb, Michael Leapman, Andrew Loblaw, Priyamvada Maitre, Jarad Martin, Giulia Marvaso, Jeff M. Michalski, Vedang Murthy, Himanshu Nagar, Paul L. Nguyen, Piet Ost, Angela U. Pathmanathan, Darren M.C. Poon, Martin Andreas Røder, Paul Sargos, Nina-Sophie Schmidt-Hegemann, Steven Seyedin, Shankar Siva, Catherine S. Spina, Simon K.B. Spohn, Daniel E. Spratt, John Nicholas Staffurth, Phuoc T. Tran, Neha Vapiwala, Constantinos Zamboglou, Nicholas G. Zaorsky, Thomas Zilli, Alison Tree, Tyler M. Seibert

## Abstract

**Purpose/Objective:** Genitourinary (GU) adverse events (AEs) are common during and after pelvic radiation therapy (RT) for prostate cancer and can substantially impact quality of life. We convened an international committee to establish consensus in the prevention, mitigation, and management of radiation-related acute and late GU AEs, as there are no relevant evidence-based consensus guidelines to inform treating providers.

**Materials/Methods:** A systematic evidence review focused on mitigation and management of radiation-related acute and late GU AEs was performed in PubMed, Embase and Cochrane. The following topics were addressed: management of acute GU AEs in the intact and post-operative settings; RT techniques; bladder outlet obstruction procedures; and indications for urology referral or hyperbaric oxygen therapy (HBO). Evidence-based consensus recommendations were developed using a Delphi process. We highlight the current state of evidence and evidence gaps worthy of future study.

**Results:** Consensus was reached for 31 key questions. For management of lower urinary tract symptoms (LUTS), most evidence comes from trials in patients without cancer and not undergoing RT. A consensus algorithm for medical management of acute GU AEs was developed with the following highlights: (a) alpha blockers as 1st-line for obstructive symptoms in the intact setting, (b) anti-spasmodics as 1st -line for irritative symptoms in the intact setting, and (c) anti-spasmodics as 1st -line in the post-operative setting. The consensus algorithm provides an ordered list of medications to offer if 1st -line options afford inadequate relief. For RT fractionation, randomized clinical trial (RCT) data are available. 40% of panelists rarely or never use standard fractionation over moderate hypofractionation for patients with baseline LUTS, but most consider moderate hypofractionation over SBRT for AUA IPSS > 15. For patients with severe obstructive LUTS (most commonly AUA IPSS >20), the panel recommends a prophylactic bladder outlet obstruction procedure and, if obstructive symptoms improve, consideration of moderate hypofractionation or SBRT, based on retrospective data. There is one RCT supporting use of HBO for late radiation cystitis.

**Conclusions:** The consensus guideline synthesizes available evidence and expert opinion across key clinical decision points to provide practical guidance in the prevention, mitigation, and management of radiation-related acute and late GU AEs in prostate cancer RT. Envisioned as a living document with periodic updates, this guideline serves as a resource for practicing radiation oncologists by outlining expert-derived consensus recommendations of evidence-based care in areas where high-quality data is limited.

## Introduction

Approximately 1.5 million men each year are diagnosed with prostate cancer worldwide, and many of them will undergo treatment with radiotherapy (RT) either in the definitive or post-operative setting^1,2^. Modern RT has excellent oncologic outcomes, but some patients experience short- and/or long-term genitourinary (GU) adverse events that can impact quality of life^3,4^. While over 30% of patients experience acute grade ≥2 GU adverse events and ≥15% of patients experience grade ≥2 late GU adverse events^5^, their management remains highly variable in practice and is supported by a limited and fragmented evidence base^6^.

In contrast to other domains in prostate cancer, such as oncologic outcomes and gastrointestinal safety, there is a notable absence of prospective, radiation-specific data to guide prevention and management of GU adverse events. As a result, clinicians frequently rely on extrapolation from non-radiation populations (for example, benign prostatic hyperplasia or overactive bladder), small retrospective series, or individual experience. This has led to substantial heterogeneity in care delivery, uncertainty in clinical decision-making and missed opportunities to optimize supportive care.

To address this gap, we convened the GU Adverse events Management (GUAM) working group to establish an international consensus guideline for best practices to prevent, mitigate, and manage acute and late GU adverse events that can occur in patients undergoing RT for prostate cancer. Using a modified Delphi process informed by a structured systematic review, we aimed to: (1) define areas of agreement and disagreement in current practice; (2) provide pragmatic, clinically implementable guidance where evidence is limited; and (3) identify critical knowledge gaps to inform future prospective investigation.

This work intentionally integrates available data with expert consensus to provide a transparent framework for decision-making in an area where randomized data are scarce. In doing so, this effort seeks not only to guide current practice, but also to establish a foundation for more rigorous, evidence-based management of GU adverse events in patients undergoing prostate external beam RT.

## Methods

### Guideline Development Process

#### Working Group Composition

The GUAM working group consists of a multi-disciplinary team (n=48) representing 15 countries from North America (n=20), Europe (n=22), Asia (n=3), and Australia (n=3) with expertise as radiation (n=34), clinical (n=7), and urologic oncologists (n=7).

#### Consensus Development

A three-round modified Delphi approach was used to reach consensus for key questions through anonymized, iterative surveys conducted via REDCap between May 2025 and February 2026^7–9^. Three authors first conducted a systematic literature review to develop the questionnaires. In round 1, the working group anonymously completed a web-based questionnaire to understand current practices and opinions. In round 2, working group members were provided evidence summaries from the systematic literature review and were asked to reconsider questions that did not reach consensus in round 1. In round 3, working group members were asked to reconsider questions that did not reach consensus from the previous rounds. Consensus was defined as ≥75% agreement^10^.

Recommendations that did not reach consensus were removed or revised between rounds. Recommendations were formulated by integrating available evidence with panel consensus; where evidence was limited, consensus thresholds were used to guide inclusion. The full questionnaires are available in the Supplemental Materials. Answers were evaluated for percent agreement for all three rounds. Working group members were instructed to answer all questions. All working group members answered round 1; over 90% answered rounds 2 and 3.

#### Evidence Review

The Preferred Reporting Items for Systematic Reviews and Meta-Analyses (PRISMA) guidelines were followed. Four databases were searched from inception to March 31, 2024: PubMed, Cochrane, Embase, and Web of Science. The reference sections of all studies that were eventually selected for full-text review were examined for additional studies. After removal of duplicates, screening of abstracts, and screening of full texts, 183 articles remained for data extraction. We repeated the search post-hoc through May 30, 2026 and did not find any new studies that would likely impact current referrals.

The Population, Intervention, Comparator, Outcome, and Study (PICOS) Design method was used to define literature inclusion criteria. Initially, we included studies of patients with localized prostate cancer undergoing definitive or postoperative RT. Secondarily, we selectively expanded the scope to include studies of men undergoing pharmacologic treatment for lower urinary tract symptoms due to benign prostatic hyperplasia or overactive bladder syndrome. The outcome of interest was acute/late genitourinary adverse events. Allowable publication types included randomized controlled trials (RCTs), prospective nonrandomized studies, meta-analyses, and retrospective studies. Universal exclusion criteria were: preclinical and non-human studies; publication types such as abstract only, case reports, comments, or editorials; and study types such as health economics or cost analysis studies. For specific sub-questions where limited data were available, expert opinion was relied on to support recommendations.

#### Scope of Guideline

From the systematic review of the literature, key questions were developed in the following topics: management of acute GU adverse events in the intact and post-operative settings; RT techniques; bladder outlet obstruction procedures; and indications for urology referral or hyperbaric oxygen therapy (HBO). The intact setting is defined as RT to the prostate gland +/− regional nodes as upfront management while the post-operative setting is defined as receiving RT after radical prostatectomy, targeting the prostate bed +/− regional nodes. The scope of this guideline is to provide practical guidance for management of acute and late genitourinary adverse events during and after prostate RT, with emphasis on acute events, which are more common. There are several important questions in the management of adverse effects of patients undergoing RT for prostate cancer that are outside the scope of this guideline, including management of acute and late gastrointestinal (GI) and sexual side effects. This guideline also does not specifically address management of adverse events attributed to other therapy for localized prostate cancer, including focal therapy, radical prostatectomy, and systemic therapy, such as androgen deprivation therapy (ADT).

#### Guideline Review & Approval

The guideline was reviewed by [xxx] official peer reviewers and revised accordingly.

#### Guideline Updating

Envisioned as a living document, the GUAM working group will conduct periodic reviews of the emerging literature to ensure this guideline remains a timely, relevant resource for practicing radiation oncologists, urologists, and other healthcare team members.

## Results – Key Questions & Recommendation

### KQ1: Medical management of Acute GU Adverse Events

Most (80%) panelists recommend against starting prophylactic medications in either the intact or post-operative setting, in anticipation of worsening symptoms during treatment, for patients not bothered by baseline urinary symptoms. A consensus algorithm for medical management of acute GU adverse events was developed (Figure 1), assuming other causes for symptoms, notably a urinary tract infection, have already been ruled out. The key features of the algorithm are: (a) alpha blockers as first-line treatment for obstructive symptoms in the intact setting, (b) anti-spasmodics as first-line treatment for irritative symptoms in the intact setting, and (c) anti-spasmodics as first-line treatment in the post-operative setting. Obstructive symptoms arise from impaired bladder emptying and include urinary hesitancy, weak or slow stream, intermittent stream, straining to void, sensation of incomplete emptying, and terminal dribbling^11^. Irritative symptoms arise from abnormalities in the bladder filling/storage phase and include urinary frequency, urgency, urge incontinence, dysuria, nocturia, and bladder pain^11^. **(Table 1).**

**Figure 1.**
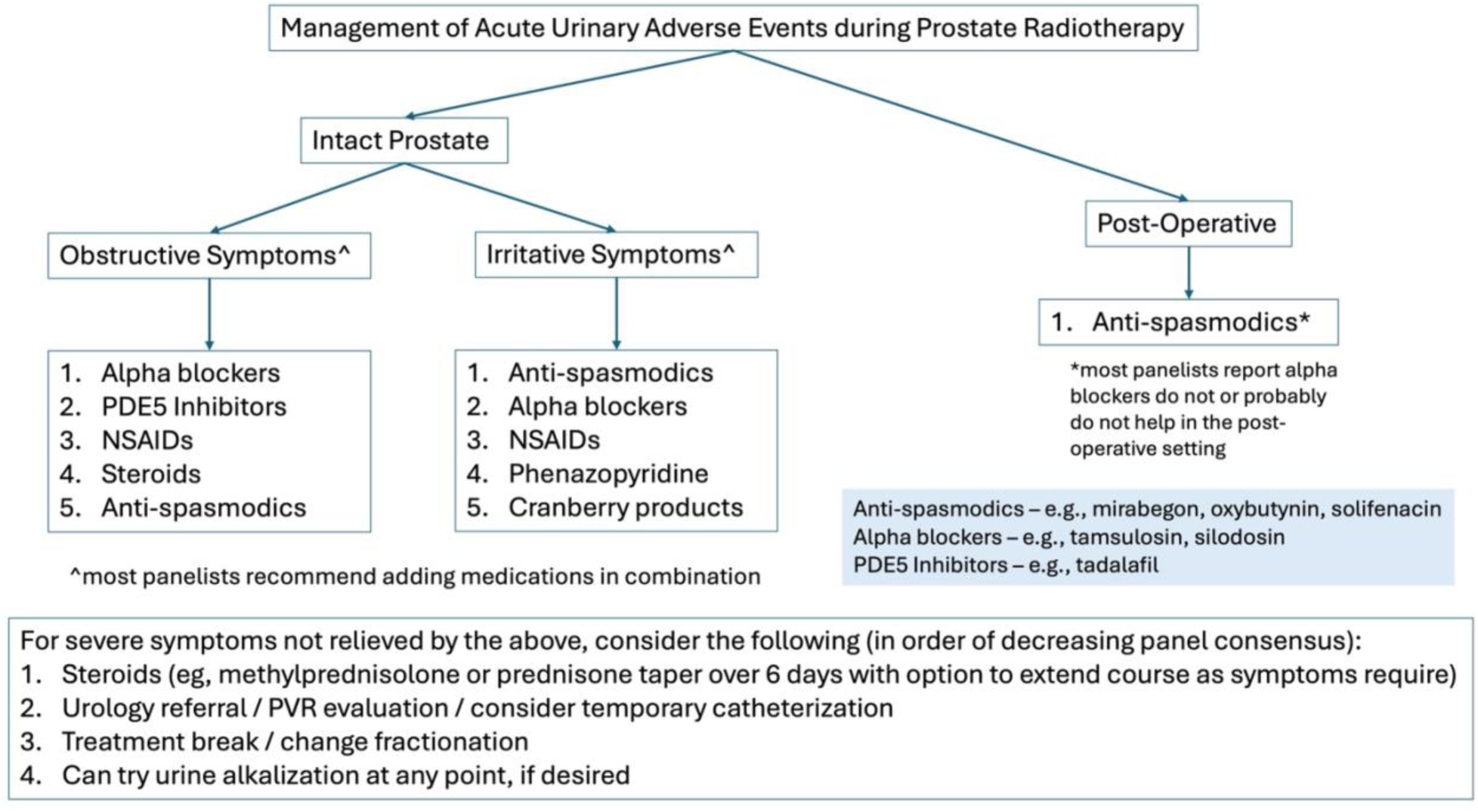
GUAM working group algorithm for management of acute urinary adverse events during and after prostate RT. PDE5: phosphodiesterase 5. NSAIDs: non-steroidal anti-inflammatory drugs. PVR: post-void residual.

**Table 1.** Obstructive vs. Irritative Urinary Symptoms.

|  | **Obstructive Symptoms** | **Irritative Symptoms** |
| --- | --- | --- |
| **Definition** | Arise from impaired bladder emptying | Arise from abnormalities in the bladder filling/storage phase |
| **Phase** | Voiding phase | Storage/filling phase |
| **Examples** | 1. Urinary hesitancy | 1. Urinary frequency |
|  | 2. Weak or slow stream | 2. Urgency |
|  | 3. Intermittent stream | 3. Urge incontinence |
|  | 4. Straining to void | 4. Dysuria |
|  | 5. Sensation of incomplete emptying | 5. Nocturia |
|  | 6. Terminal dribbling | 6. Bladder pain |

Most (74%) panelists believe alpha-blockers do not or probably do not help in the post-operative setting. The consensus algorithm provides an ordered list of medications to offer if first-line options afford inadequate relief. Supplemental Table 1 provides a summary of medication names and mechanisms of action, starting dose, maximum dose, and key contra-indications.

Several medications fall into the category of anti-spasmodics. 15% of panelists had no preference on which anti-spasmodic to be used. Among those with a preference, 45%, 28%, and 21% preferred mirabegron, solifenacin, and oxybutynin, respectively. If the first medication provided inadequate relief, 80% of panelists recommended adding a second class of medication in combination (rather than discontinuing the first). Lastly, the GUAM panel recommends several options in the case of persistent symptoms not adequately relieved by the medications in the consensus algorithm.

#### Evidence to Support KQ1

##### Evidence for Alpha Blockers (e.g., tamsulosin, silodosin) for obstructive symptoms

There have been 5 randomized controlled Trials (RCTs) for use of preventative alpha blockers in the setting of prostate brachytherapy; all showed some degree of improved obstructive lower urinary tract symptoms (LUTS), measured by decrease in International Prostate Symptom Score (IPSS) with the use of these medications^12–16^. A single RCT in the setting of prostate external beam radiation therapy (EBRT) (n=108) found that six months of tamsulosin (starting on day 1 of RT) reduced acute urinary retention compared to no intervention^17^. A single arm prospective trial (n=29) of tamsulosin use in the setting of EBRT found that 50% of patients had progressive LUTS on 0.4 mg tamsulosin despite initial response and of those, 75% had a durable response with doubling of dose to 0.8 mg^18^.

Another RCT (n=148) found that starting silodosin prophylactically at start of RT (compared to starting silodosin only at onset of LUTS) did not improve patient-reported LUTS measured by IPSS^19^.

Several RCTs compared silodosin to tamsulosin or placebo for patients with LUTS from benign prostatic hyperplasia (no radiation was given). Silodosin and tamsulosin both improved patient-reported LUTS (IPSS total score, IPSS voiding subscore, IPSS storage subscore and quality of life) more than placebo and neither medication was proven superior to the other^20,21^.

Several retrospective series and single-arm studies have confirmed that alpha blockers are generally well tolerated with only mild side effects^22–27^. Besides medication management, our working group recommends consideration of behavior changes, such as reducing evening fluid intake, if appropriate.

##### Evidence for Phosphodiesterase type 5 inhibitors (PDE5-Is) for obstructive symptoms

Phosphodiesterase type 5 inhibitors (PDE5-Is), specifically tadalafil 5 mg once daily, are supported by robust evidence from RCTs for the management of LUTS due to benign prostatic hyperplasia (BPH). The most comprehensive meta-analyses and systematic reviews have included up to 16 RCTs, with over 4,000 participants, demonstrating that PDE5-Is yield a modest but statistically significant improvement in IPSS compared to placebo (mean difference approximately 1.9–2.8 points) and similar efficacy to alpha-blockers for symptom relief. Combination therapy with alpha blockers and PDE5-Is provides incremental benefit in IPSS reduction, but with increased side effects, such as headache, myalgia, and dizziness^28^. The evidence is primarily short-term (up to 12 weeks), and long-term data on symptom control are lacking. Importantly, there are no RCTs or single-arm studies evaluating PDE5-Is for LUTS specifically in the context of RT; all available data pertain to BPH-related LUTS, and no guideline or trial addresses post-radiation LUTS management with PDE5-Is^29–33^.

##### Evidence for therapies for irritative urinary symptoms

###### Anti-spasmodic (e.g., oxybutynin, mirabegron)

A single RCT in the setting of prostate brachytherapy (n=218) found that the addition of mirabegron (mechanism of action: beta-3 agonist) to tamsulosin improved overactive bladder symptom score and 24-hour urinary frequency but did not improve voided volume per micturition compared to tamsulosin alone^34^. There are no randomized trials specifically evaluating anti-spasmodic therapy for urinary adverse events management in the setting of prostate EBRT. Interestingly, a recent RCT (n=88) found oxybutynin (mechanism of action: anti-cholinergic) reduced hot flashes compared to placebo and is well tolerated, with dry mouth as most common side effect^35^.

However, there are >100 RCTs supporting the use of anti-spasmodic medications (oxybutynin, mirabegron, vibegron) for patients with overactive bladder syndrome. Two meta-analyses (n=86 trials, n=60 trials) found oxybutynin, tolterodine, and solifenacin were all significantly better than placebo in improvement of urinary urgency, frequency, incontinence, and nocturia^36,37^. Solifenacin was found most effective for improving urinary frequency, incontinence, and nocturia, while oxybutynin was most effective for increasing voided volume per micturition. Anticholinergic drugs are well tolerated but likely to cause dry mouth and constipation; lower doses or extended-release preparations are better tolerated.

A meta-analysis of 64 RCTs (n=46,666) demonstrated that mirabegron was superior to placebo in relief of overactive bladder symptoms and similar to a range of common anticholinergics (including oxybutynin, tolterodine, solifenacin) with significantly less anti-cholinergic side effects including dry mouth and constipation^38^. Given the potential for anti-cholinergic medications to exacerbate age-related memory-issues^39^ (not seen with mirabegron), we recommend exercising caution in the case of long-term use and in patients with underlying issues.

###### Phenazopyridine

Phenazopyridine was initially marketed in 1938 and grandfathered into FDA for approval. There is limited prospective data (and no RCTs) in the setting of RT.

A single double-blinded RCT (n=60) found that for patients with urinary tract infections (no RT given), a single dose of phenazopyridine (given prior to initiation of antibiotic therapy) improved irritative symptoms compared to placebo^40^.

A single retrospective series reports that extended use (>14 days) of phenazopyridine during RT was safe and well tolerated^41^.

###### Cranberry Capsules

There are 3 randomized, placebo-controlled trials for use of cranberry capsules during EBRT, with conflicting results.

Two positive trials (n=924, n=41) demonstrate that cranberry capsule use significantly improved urinary discomfort and urinary stream strength with reduction in antibiotic use compared to placebo^42,43^.

A third study (n=108) reported cranberry capsules were not significantly better than placebo in management of radiation cystitis^44^.

###### Urinary alkalization

There have been no RCTs evaluating urinary alkalization in the setting of prostate RT^45^.

A single trial (n=59) found that for female patients with overactive bladder syndrome (no radiation given) urinary alkalization with oral sodium bicarbonate improved symptoms and urinary quality of life similarly to treatment with solifenacin^46^.

Several single arm trials confirm that for patients with hyperactive bladder symptoms (no radiation given) urinary alkalization improves symptoms and is well-tolerated^47,48^.

### KQ 2 – Radiation Techniques and GU Adverse Events

We sought consensus on several practical aspects of a patient’s radiation treatment plan that could contribute to urinary adverse events, including fractionation, bladder filling, and dose constraints. While consensus was reached for fractionation recommendations and bladder filling, we highlight an evidence gap for dose constraints correlated with acute or late urinary adverse events.

#### Evidence to Support KQ2

##### Fractionation Scheme

Given the various fractionation schemes available for RT to the intact prostate, the GUAM panel provides recommendations regarding selection of the appropriate fractionation scheme for a patient based on pre-treatment urinary symptoms. A full breakdown of panelists’ fractionation preferences based on IPSS is shown in Figure 2. Most (76%) panelists favor moderate hypofractionation over SBRT for IPSS 15-19. Consensus was not reached regarding when to recommend standard fractionation. Of note, 41% of panelists *never* recommend standard fractionation (around 2 Gy per fraction) over moderate hypofractionation (around 3 Gy per fraction) due to a high IPSS.

**Figure 2.**
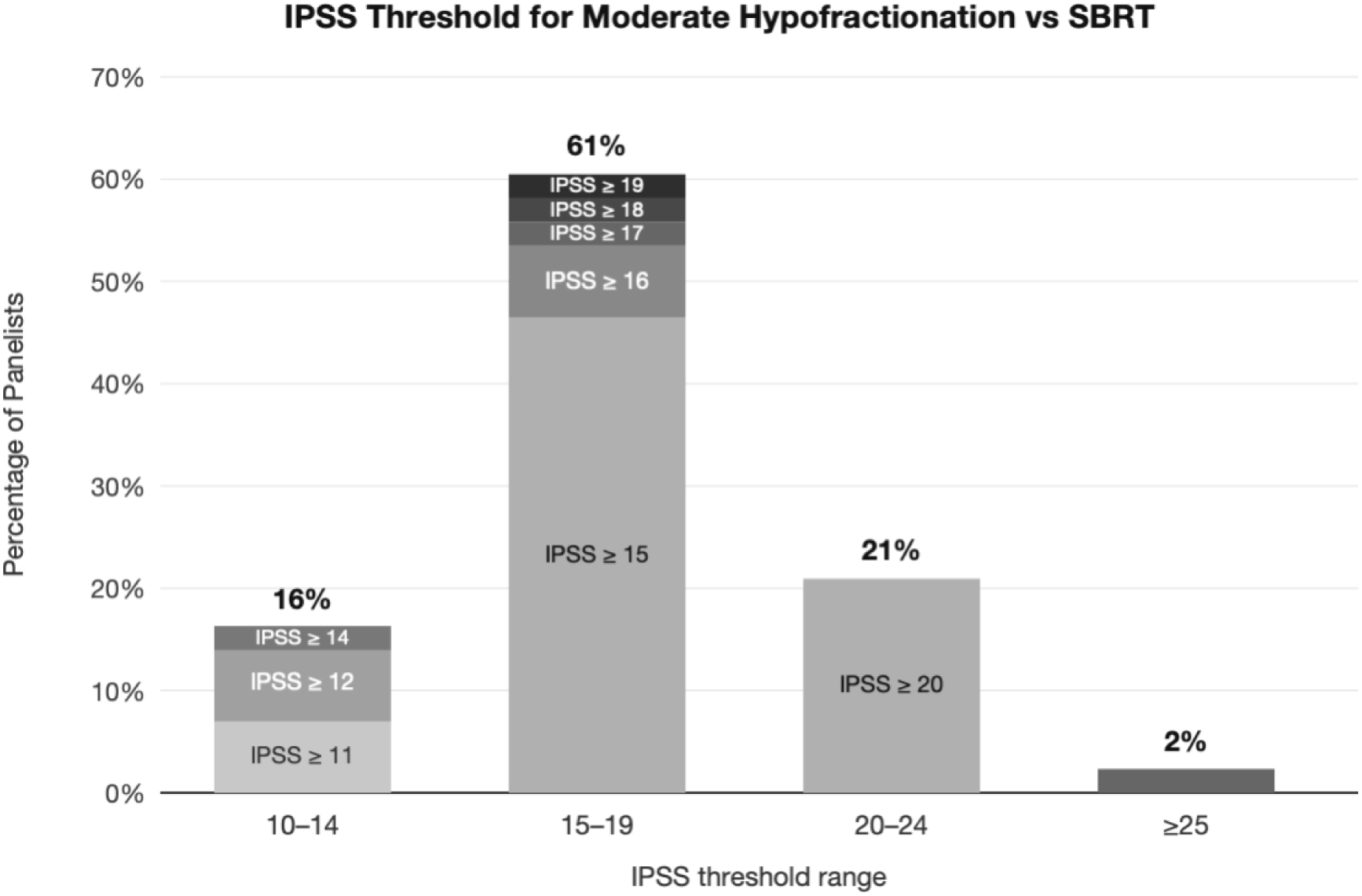
IPSS threshold at which moderate hypofractionation is generally favored over SBRT. The most common IPSS threshold chosen was 15 (60% of panelists). Most (76%) panelists favor moderate hypofractionation for IPSS 15-19, and nearly all (97%) favor moderate hypofractionation over SBRT if IPSS ≥20. Different shades of gray indicate distinct IPSS thresholds recommended by panelists.

##### Evidence for fractionation based on LUTS

Three RCTs (CHHiP, RTOG 0415, PROFIT) compared conventional and moderate hypofractionation for prostate EBRT^49–51^. They did not exclude patients based on urinary symptoms. All three studies show no difference in acute urinary adverse events between conventional and moderate hypofractionation.

Two studies (CHHiP and PROFIT) reported similar rates (∼20%) of late grade 2 urinary adverse events between both groups. However, RTOG 0415 reported significant increase in late GU grade 2+ adverse events with moderate hypofractionation compared to conventional fractionation. Recently, the HYDRA meta-analysis demonstrated that isodose moderate hypofractionation showed no significant increase in late GU adverse events compared to conventional fractionation (5-year late grade ≥2 GU adverse events 23.9% vs. 22.3%. In contrast, dose-escalated moderate hypofractionation was associated with significantly higher late grade ≥3 GU adverse events (7.2% vs. 4.9) without significant differences in 5-year late grade ≥2 adverse events^52^.

Retrospective studies report that higher pre-treatment IPSS is associated with higher incidence of grade ≥2 acute and late GU adverse events for both EBRT and brachytherapy^53,54^.

There are no prospective trials specifically evaluating safety and tolerability of SBRT in patients with high baseline IPSS. A single-institution, retrospective study (n=72) reported a 44% late grade 2 GU adverse events rate in patients with IPSS ≥15 treated with SBRT^55^. A second, single-institution, retrospective study (n=53) reported decreased IPSS from baseline at 3 months after SBRT that remained stable at 36 months post-treatment in patients with IPSS ≥15 treated with SBRT^56^. PACE-B, an RCT evaluating SBRT versus moderate hypofractionation (n=874) included patients with moderate to severe baseline LUTS – approximately 33% of patients had IPSS between 8-19 (moderate symptoms), and an additional 5% had IPSS ≥20 (severe symptoms). There was a modest increase in cumulative risk of grade ≥2 GU adverse events at 5 years in the SBRT arm (26.9%) versus moderate hypofractionation arm (18.2%)^57,58^. A post-hoc exploratory analysis of PACE-B found the incidence of grade ≥2 adverse events at 2 years was 8% in patients with a baseline IPSS of ≤11, compared with 27% in those with a baseline IPSS of >11^59^. Early safety results of PACE-C, an RCT evaluating SBRT versus moderate hypofractionation (n=1208), report that acute physician-reported CTCAE grade ≥2 GU adverse events were significantly increased in patients with baseline LUTS compared with those without^60^. Furthermore, PRIME is an ongoing RCT evaluating the non-inferiority of SBRT versus moderate hypofractionation in patients with high risk and node positive prostate cancer but does exclude patients with prior history of transurethral resection of prostate (TURP), severe urinary symptoms despite 6 months of ADT (defined as IPSS >15), or known stricture^61^.

##### Bladder Filling

Regarding bladder filling instructions, 85% of panelists recommend instructing patients to have a full bladder for simulation and treatment. 10% of panelists instruct patients to empty their bladder prior to simulation and treatment, with the remaining 5% of panelists providing no bladder fill instructions to patients.

###### Evidence for Bladder Filling Protocol during RT

There are no prospective studies demonstrating a patient benefit to bladder filling instructions. Retrospective, non-randomized, single-institution studies report that bladder empty protocols reduced absolute variation in bladder volume during treatment, had minimal impact on treatment-related adverse events, and had non-inferior biochemical progression free survival, GI safety, and GU safety^62–65^.

Two ongoing RCTs are directly comparing full versus empty bladder filling instructions – BEFORE: Bladder Full OR Empty for pelvic radiation therapy (clinicaltrials.gov ID: NCT06651697) and RELIEF: Randomized EvaLuation of the Impact of Empty versus Full Bladder (clinicaltrials.gov ID: NCT06037863).

##### Dose Constraints

There was a lack of consensus for the most important dose constraints for preventing urinary adverse events, reflecting the evidence gap in the current literature.

###### Dose Constraints

Characterizing a reliable relationship between dose/volume parameters and acute/late urinary adverse events remains elusive^66^. The only QUANTEC paper that does not provide any dose-volume guidelines to mitigate risk of adverse events is the one on urinary bladder, due to a lack of clear association^67^. We found insufficient evidence to support any specific set of dose goals for reducing GU AEs. We endorse the ALARA principle and call for prospective studies of how dosimetry can affect GU AEs. With the advent of MRI in RT planning, there is increasing interest in evaluating dose to the urethra as well as substructures of the bladder as predictors of urinary adverse events^68^.

###### Evidence for Urethral Dose Constraints

While data on urethral dose constraints from conventional fractionation is inadequate because of difficulty visualizing the urethra with traditional CT imaging, MRI target volume localization allows for assessment of urethral dose^69^.

The investigators of the FLAME trial published a dose-effect analysis of FLAME with suggested urinary dose goal of D0.01cc ≤105% that is now being used prospectively in hypo-FLAME^57,70,71^.

Additionally, the HypoFocal-SBRT trial is using both urethra D0.01 cc < 40Gy and D50% < 36 Gy as it is currently still unclear which urethral dose-volume constraints are clinically useful^72^.

A prospective study of 110 physician contours on MRI found severe variation across physicians and poor agreement with the expert consensus reference standard^73^. These findings raise questions about the reliability of previously reported urethra dose associations with adverse events and doubts about the feasibility of widely implementing dose goals in general practice. Use of contouring atlases and artificial intelligence tools has shown promise in facilitating study of this question^74–76^.

###### Planning Margins

Whilst not specifically addressed by our consensus exercise, recent evidence from the MIRAGE trial has shown that smaller margins, used in that trial alongside MRI guidance, can significantly reduce GU adverse events. Acute GU adverse events (grade 2 or higher) were reduced from 43.4% (CT guidance, 4 mm margins) to 24.4% (MRI guidance, 2 mm margins). Long-term grade ≥2 GU adverse events were reduced from 51% (4 mm margins) to 27% (2 mm) margins by 2 years^77^. Secondary analysis has suggested that intrafraction motion control may further reduce adverse events^78,79^.

### KQ 3- Bladder Outlet Obstruction Questions

Most (65%) panelists do not recommend requiring routine measurement of urodynamics (post void residual (PVR) and flow rate) prior to starting RT. Approximately 25% of panelists routinely get PVR/flow rate prior to RT and most of these panelists recommend this assessment even if irritative symptoms predominate over obstructive symptoms.

85% of panelists recommend consideration of a bladder outlet obstruction procedure prior to RT in the setting of severe LUTS. However, almost all panelists (95%) agree that a bladder outlet obstruction procedure should not be considered if irritative symptoms predominate over obstructive symptoms. While most panelists use a numerical threshold on the IPSS instrument as an indicator of need for bladder outlet obstruction procedure (Figure 3a), some providers also incorporate urinary quality of life assessments from patients or data from urinary dynamics assessment. For panelists using PVR/flow rate to determine need for bladder outlet obstruction procedure, common thresholds include flow rate Qmax <10 mL/second and/or PVR values between 100-300 mL.

**Figure 3.**
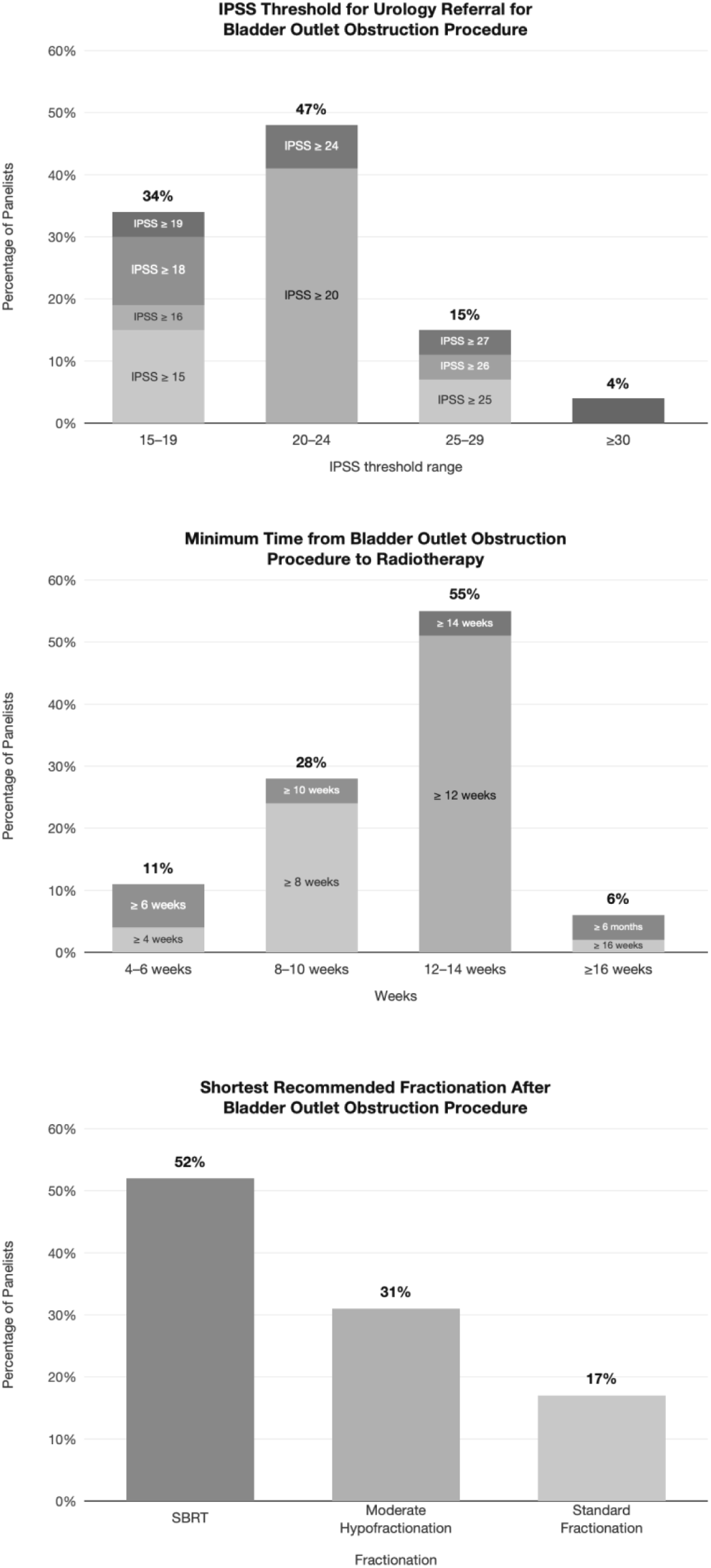
Panelists’ recommendations regarding bladder outlet obstruction procedures (e.g., transurethral resection of prostate, or TURP). Top: IPSS threshold at which to generally refer to urology for consideration of a procedure. Most (74%, just shy of the *a priori* consensus threshold) panelists would typically refer a patient with IPSS ≥20 for consideration of a procedure. Middle: Minimum time to allow for post-procedure recovery before initiating RT. Nearly all (94%) panelists recommend at least 8 weeks to recover, and most (66%) recommend ≥12 weeks. Bottom: Shortest fractionation typically recommended in patients who have had a procedure. Only 17% of panelists insist on standard fractionation after a successful procedure, while most (52%) consider SBRT a safe option for patients who have minimal urinary dysfunction after recovery from the procedure. Different shades of gray indicate distinct thresholds recommended by panelists for each question.

If a bladder outlet obstruction procedure is performed, 75% of panelists recommend waiting at least 8 weeks before starting RT, with 50% of panelists recommend waiting 12 weeks before starting RT, noting that systemic therapy may be started earlier as clinically indicated (Figure 3b). Regarding fractionation scheme after bladder outlet obstruction procedure, most panelists recommend either moderate hypofractionation or choosing fractionation scheme based on post-procedure LUTS severity. Only 15% of panelists recommend standard fractionation for these patients. Additionally, 52% of panelists agree that SBRT is acceptable for patients who underwent bladder outlet obstruction procedures for severe LUTS and now have minimal obstructive urinary symptoms (Figure 3c).

Almost all panelists recommend bladder outlet obstruction procedure for persistent severe LUTS after RT. 50% of those panelists reporting the adverse events rate is not much worse compared to bladder outlet obstruction procedure in the absence of prior RT, whereas the 2019 American Urological Association guideline has a moderate recommendation (with low-quality available evidence) that patients undergoing transurethral should be advised of a higher risk of incontinence^80^.

#### Evidence to Support KQ3

##### Evidence for Bladder Outlet Obstruction Procedure after RT

There are no prospective trials evaluating bladder outlet obstruction procedures after RT.

Several retrospective studies reported that bladder outlet obstruction procedure after RT (brachytherapy or EBRT) was associated with new-onset urinary incontinence in up to 27% of patients^81,82^.

### KQ 4 - Urology Referral

#### Evidence to Support KQ4

There are no prospective data to guide the decision for when to refer patients to urology for acute and late side effects, in either the intact or post-operative setting.

Our panelists provide recommendations for when to refer patients undergoing RT for intact prostate to urology. The most common reason for referral in the acute setting is for patients with persistent/severe LUTS after all medication management options are exhausted. Following this recommendation, panelists also endorse consideration of urology referral for obstructive symptoms, persistent bleeding, urinary retention and new catheterization. Similarly, panelists recommend urology referral for management of late GU adverse events (in order of decreasing consensus recommendation): persistent hematuria, exhausted medication options for LUTS, obstruction, retention, and new incontinence.

In the post-prostatectomy setting, our panelists recommend urology referral for the following acute GU adverse events: exhausted medication options, obstruction/stricture, hematuria, incontinence, and need for catheterization. Similarly, the GUAM panel recommends urology referral for the following late GU adverse events: bleeding, stricture, incontinence, exhausted all medication options, and catheterization. We highlight the differences in referral reasons in the intact setting versus post-prostatectomy – panelists recommended referral for bleeding, strictures, and incontinence more commonly in the post-prostatectomy setting, mirroring which GU adverse event are most common after surgery.

### KQ 5 – Hyperbaric Oxygen (HBO) Therapy Referral

Panelists reported different thresholds for referral for HBO in the late setting, but common themes included needing a cystoscopy before to establish radiation cystitis and to only refer if persistent hematuria after optimization of medical management.

#### Evidence to Support KQ5

##### Evidence for Hyperbaric Oxygen Therapy for Radiation Cystitis

A solitary RCT, RICT-ART, (n=87) demonstrated that HBO significantly improves symptoms of late radiation urinary adverse events as measured by patient-reported questionnaire, EPIC, as compared to standard care and is safe and can be well tolerated^83^.

## Limitations

While panelists are experienced in treatment of prostate cancer and managing adverse events after RT, they do not represent a random sample of all physicians or even of experts. There were few panelists from Asia and none from Africa or South America. Medications recommended may be variably available in different practice settings. Possible geographic or cultural variations in patient preferences have not been well studied and are not addressed here. Additionally, this does not include urinary assessment and recommendations for patients being treated with brachytherapy boost.

## Conclusion & Future Directions

The consensus guideline synthesizes available evidence and expert opinion across key clinical decision points to provide practical guidance in the prevention, mitigation, and management of radiation-related acute and late GU adverse events during and after prostate cancer RT. We provide recommendations for the five following key questions: (1) medication management of acute GU adverse effects in both the intact prostate and post-prostatectomy setting; (2) management of bladder outlet obstruction prior and after RT; (3) techniques in RT planning and delivery to mitigate urinary adverse events; (4) when to refer patients to urology; and, (5) the role of HBO. In the systematic review, we provide evidence for our recommendations.

Through this systematic review, we uncovered evidence gaps and lack of prospective data to direct many of these important decisions practicing radiation oncologists make daily. Because data to guide these are scarce, we formed an international working group to derive expert consensus guidelines. We posit the primary contribution of this work is not the introduction of new therapeutic strategies, but rather the systematic identification and structuring of clinical decision-making in an area characterized by uncertainty. By delineating where consensus exists, where it does not, and where evidence is lacking, this guideline provides a practical framework for clinicians while simultaneously defining a research agenda for the field. Thus, we call for more RCTs to guide care to improve patients’ lives. We acknowledge that treatment-related adverse events are multi-factorial, including history of prior LUTS, RT technique and dose. In some cases, other cancer-directed local therapies like surgery or focal therapy play a role. Even ADT may impact urinary adverse event rates, as seen in RCTs of RT with or without ADT^3,84^. There are ongoing investigator-initiated trials to address (and mitigate) urinary adverse events due to RT: BEFORE (NCT06651697)^85^, RadTARGET (NCT06990542)^86^, RELIEF (NCT06037863), and DESTINATION 2 (NCT06638541)^87,88^.

Beyond improving management of treatment related adverse events, the GUAM working group recommends increased focus on the causes of adverse events and their risk factors (e.g., genetic susceptibility). Envisioned as a living document with periodic updates, this guideline serves as a resource for practicing radiation oncologists in the management and mitigation of GU adverse events during and after RT for prostate cancer.

## Data Availability

All data produced in the present study are available upon reasonable request to the authors

## Author Contributions

Anna M Dornisch is the first author. Alison Tree and Tyler M. Seibert are co-senior authors. Tyler M. Seibert is the corresponding author.

## Supplemental

**Supplemental Table 1:**
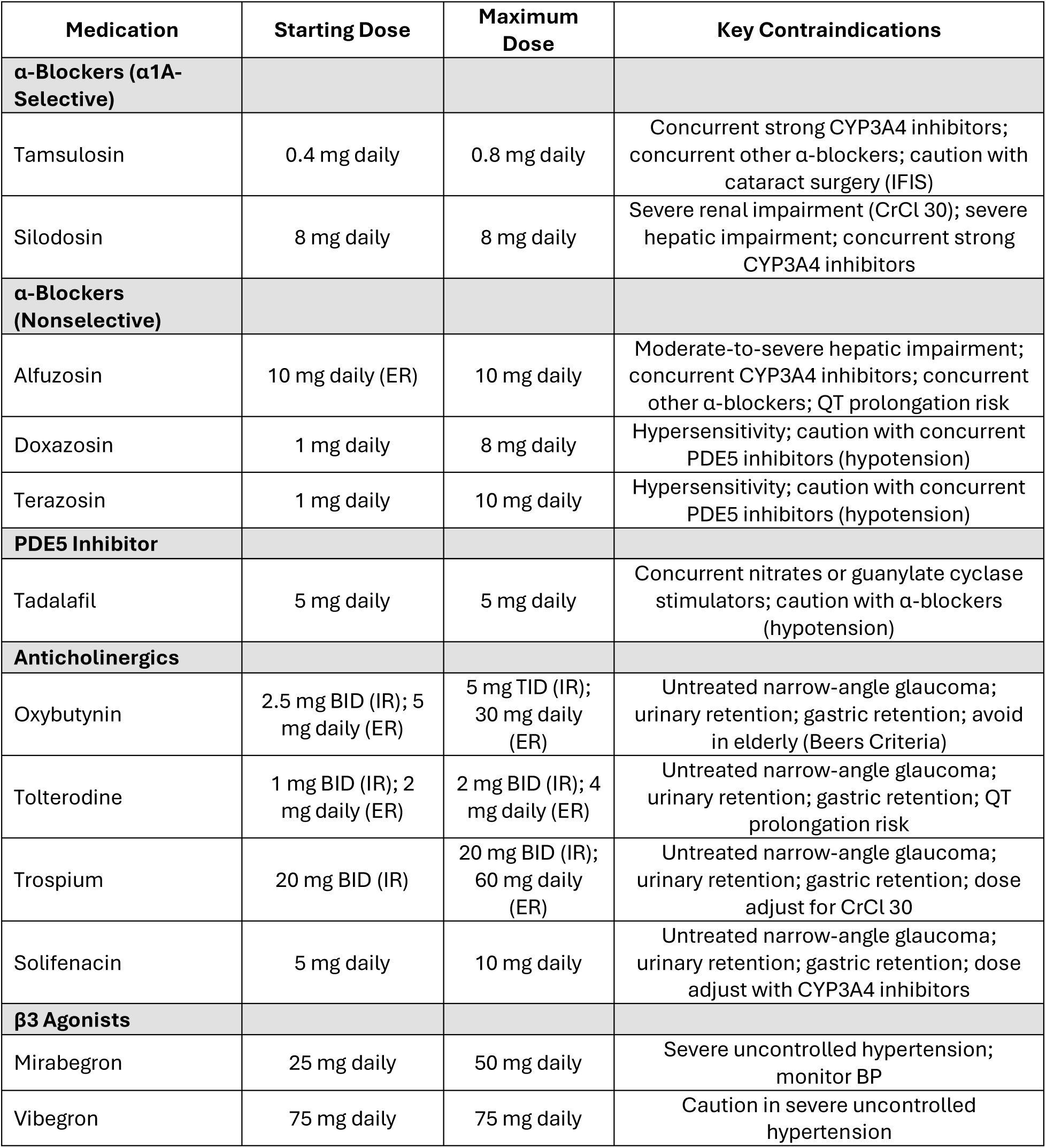
Summary of commonly used medications, including recommended starting doses, maximum doses, and key contraindications relevant to clinical prescribing considerations.

